# Access to community-based eye services in Meru, Kenya: a cross-sectional equity analysis

**DOI:** 10.1101/2024.02.23.24303185

**Authors:** Luke Allen, Sarah Karanja, Michael Gichangi, Cosmas Bunywera, Hillary Rono, David Macleod, Min Jung Kim, Malebogo Tlhajoane, Matthew J. Burton, Jacqueline Ramke, Nigel M. Bolster, Andrew Bastawrous

## Abstract

**Background:** Over 80% of blindness in Kenya is due to curable or preventable causes, with an estimated 7.5 million Kenyans in need of quality eye care services. Embedding sociodemographic data collection into the national eye screening programme could help identify the groups facing systematic barriers to care. We aimed to determine the sociodemographic characteristics that are associated with access among patients diagnosed with an eye problem and referred for treatment in the national eye screening programme.

**Method:** We used an embedded, pragmatic, cross-sectional study design. A list of sociodemographic questions was developed with input from researchers, community members, policymakers, and programme implementers. After five rounds of iteration, the final sociodemographic question set included the following domains: age, gender, religion, marital status, disability, education, occupation, income, housing, assets, and health insurance. These were integrated into an app that is used to screen, refer, and check-in (register) participants within a major eye screening programme. We gathered data from 4,240 people who screened positive during community screening and were referred to a local outreach treatment clinic in Meru County. We used logistic regression to identify groups for whom services were inaccessible.

**Findings:** Only 46% of those who were referred to local treatment outreach clinics were able to access care. In our fully adjusted model, at the 0.05 level there were no statistically significant differences in the odds of attendance within the domains of disability, health insurance, housing, income, or religion. Strong evidence (p<0.001) was found of an association between access and age, gender, and occupation, with males, younger adults, and those working in sales, services and manual jobs being the least likely to access care.

**Conclusions:** Less than half of those identified with an eye need and referred to free local clinics were able to access care in Meru. Younger people are being left behind, with less than a third of those aged 18-44 receiving care. Future work should explore the barriers and potential solutions to equitably improve access to care for this group.

## Introduction

More than one billion people currently live with preventable or untreated visual impairment, and over 90% of these cases are easily treatable with highly cost-effective interventions like spectacles and cataract surgery.^7^ The vast majority of people with untreated eye conditions live in low- or middle-income countries, and within these countries marginalised groups are often disproportionately affected.^7–9^ Extending access to eye services is a global health priority that aligns with both the principles of proportionate universalism^1^ and Primary Health Care: an approach to health that prioritises the worst-off and seeks to advance equity and *health for all*.^2^

An estimated 7.5 million people require eye health services in Kenya, but less than a quarter are able to access services.^3^ In 2022 the government launched the ‘Vision Impact Programme’ (VIP) in which community-based teams use smartphones to administer ‘tumbling E’ visual acuity assessments, using an app developed by the social enterprise Peek Vision.^4^ Those who screen positive - i.e. their visual acuity is found to fall below a predetermined threshold (<6/12 in either eye) are referred to a local outreach treatment clinic, commonly held in a primary care facility, where they receive free further assessment and care, including spectacles, eye drops, or onward referral for cataract surgery at a local hospital as required. Screeners also refer people who have a red eye or another issue upon basic visual inspection, and anyone who feels they have an eye problem, even if there are no clinical signs and their visual acuity is >6/60.

In the VIP programme’s first year, over a million people were screened and more than 150,0000 were managed at free treatment outreach clinics.^5^ Whilst this is a remarkable achievement, internal Peek data suggest that there are important issues with clinic accessibility, as less than half of those who were identified with an eye problem during community-based screening received care at their local clinic.

Access is determined by both patient and provider factors,^6^ and evidence from other countries suggests that certain groups such as females, widows, and those in rural areas - may face unique structural barriers to accessing eye care services.^7^ Currently, no sociodemographic data beyond age, gender, and language are being collected in the VIP screening programme, and these data are not currently being used to perform equity analyses. As such, any sociodemographic inequities are invisible.

Acknowledging the risk that “poorer, less advantaged segments of the population could be left behind” as countries expand access to health services in pursuit of UHC, joint WHO and World Bank guidance recommends that health programmes routinely gather data on gender, wealth, and place of residence (urban/rural) to monitor equity in effective service coverage.^8^ The recent UN Resolution on Vision, the Lancet Commission on Global Eye Health, and the Declaration of Astana all call on global health partners to analyse the equity impact of their programs across different sociodemographic populations.^9–11^ This aligns with the ‘central transformative promise’ of the Sustainable Development Goals which is to ‘leave no one behind’ and the commitment to ‘reach the furthest behind first’.^12^

Working with the Ministry of Health, a local community advisory board, the VIP programme implementing partner, and Peek Vision, we aimed to integrate a set of sociodemographic questions into the community-based screening process in Meru county and perform the first assessment of whether all sociodemographic groups are experiencing similar levels of access to primary eye care.

## Methods

### Population

The VIP programme has been designed to screen all residents aged over 18 years in ten of Kenya’s 47 counties.^13^ Working with the national director of eye services, we identified Meru county as the best place to conduct our study, based on the fact that it contains a mix of urban and rural areas, has a leadership engaged with equity-focused quality improvement, and had a screening schedule that aligned with our research timeline. Meru is a central high-altitude county on the slopes of Mount Kenya with a population of 1.55 million, most of whom live in Meru town, the seventh largest urban centre in the country. Agriculture is the main source of employment, with khat and tea being the most prevalent cash crops.

### Sociodemographic domains

We started by performing a literature review and a secondary analysis of data from a systematic review to identify the sociodemographic domains that are being used by other programmes, agencies, and researchers around the world. Full details and results are available in our published protocol.^14^ Briefly, we identified 11 broad domains that had been used or recommended in the peer-reviewed literature and UN agency reports: age, gender, residence (urban/rural), language, ethnicity/tribe/race/caste, refugee/immigrant status, marital status, religion, occupation, income, and wealth.^8–10,15–19^ We drafted response options for each domain that aligned with those used in the widely-used USAID Demographic and Health Survey (DHS) that has been used to complete more than 400 surveys in 90 countries^20,21^ and the Rapid Assessment of Avoidable Blindness (RAAB) instrument that has been used for over 300 surveys in 80 countries.^22^ This was to ensure that all ensuing data complied with international norms and were maximally useful for domestic policymakers.

Next, we set up a multi-stakeholder workshop that included representatives from Peek Vision, the implementing partner organisation (Christian Blind Mission), the Ministry of Health, and local academics with experience and expertise in sociodemographic data collection. This group adapted each of the draft domains to the Kenyan context, and adding in a housing question as an indicator of wealth.

Over the course of four hybrid workshops, we iteratively refined the list of domains and questions stems, seeking to align them with pre-existing locally collected data and ensuring that the wording accorded with cultural norms. We removed the question on tribe/ethnicity as this was considered to be potentially inflammatory. Supplementary tables 1-4 present further detail on the decisions made at each stage.

All decisions were made by consensus, and after five rounds of iteration the final list included 11 domains with between 2-8 individual response options (Table 1). Every domain also included ‘don’t know’ and ‘do not want to answer’. The draft survey instrument was translated into Kiswahili and back-translated into English to check that meaning had not been lost. The survey was piloted with laypeople using a ‘think aloud’ approach,^23^ and then in the actual screening programme with approximately 100 service users. No changes were indicated during piloting.

**Table 1:** Sociodemographic domains and response options.

| **Domain** | **Question stem** | **Response options** |
| --- | --- | --- |
| **Gender** | What is your gender? | Female |
|  |  | Male |
|  |  | Other |
| **Age** | What is your age? | 18-24 |
|  |  | 25-34 |
|  |  | 35-44 |
|  |  | 45-54 |
|  |  | 55-64 |
|  |  | 65+ |
| **Language** | What is your preferred language? | Kiswahili |
|  |  | English |
| **Marital status** | What is your marital status? | Single |
|  |  | Married |
|  |  | Divorced/separated |
|  |  | Widowed |
| **Assets** | Does your household own a bicycle, motorbike, scooter, car, or truck? | None |
|  |  | Bike or Moto or Scooter |
|  |  | Car or Tuck |
| **Disability** | Do you have any difficulty with hearing, walking, climbing steps or communicating? | No |
|  |  | Yes (one or more) |
| **Education** | What is your highest level of education? | None |
|  |  | Primary |
|  |  | Secondary |
|  |  | Post-secondary |
| **Health insurance** | Do you have health insurance? | No |
|  |  | Yes, active |
|  |  | Yes, not active |
| **Housing** | Do you have Electricity, Solar, or a Generator at home? | No |
|  |  | Yes |
| **Income** | In the last month, what was your approximate income? | KES <24,000 |
|  |  | KES 24,000-32,333 |
|  |  | KES >32,333 |
| **Occupation** | What is your occupation? | Not employed |
|  |  | Farming |
|  |  | Domestic service |
|  |  | Professional* |
|  |  | Sales & services |
|  |  | Skilled manual |
|  |  | Unskilled manual |
|  |  | Student/pupil |
| **Religion** | What is your religion? | Christian |
|  |  | Islam |
|  |  | Hindu |
|  |  | Other |

\*Note: Includes professional or manager or technician or clerical

### Screening approach

In the VIP programme, community health workers go house-to-house and assess the vision of all residents. For each participant, they enter the following demographic details into the Peek app:^24^ name, contact phone number, age, and gender. Next, they perform a ‘tumbling E’ visual acuity assessment using a smartphone. As stated above, if the participant’s vision falls below a pre-specified acuity threshold, or if they have a visible or reported subjective eye complaint (e.g. a red or painful eye), then the participant is referred to the local clinic for further assessment and treatment. At this point their preferred language is recorded. The participant is given an appointment date and is sent a follow-up reminder text message. On the day of assessment, participants are checked-in (registered) by staff using the same Peek app at the clinic. This means that Peek hold a record of all those referred and can generate a complete list of all those who have and have not been checked-in on their appointed date.

We added the extended list of sociodemographic questions to the Peek app. These questions were asked of every person who was found to have an eye problem and referred to their local treatment outreach clinic. Informed written consent to gather these additional sociodemographic data was obtained by the community health workers who performed the screening, using paper consent forms.

### Sample size

Our aim was to compare the odds of attendance between different sociodemographic subgroups (e.g. males vs females). Our community advisory group suggested that we would want to detect differences in attendance of 5-10% or more between subgroups. With a 95% confidence level and a maximally conservative proportion of 50% attendance, we calculated that we would need to have at least 1,566 people in each subgroup to have 80% power to detect a 5% difference between subgroups, or 385 people in each subgroup to detect a difference of 10%. We decided to set our sample size at 3,850 which would provide 80% power to detect differences of 10% between groups that contain at least 10% of the overall population, while still providing power to detect a difference of 5% in subgroups that make up 40% of the population. We deemed that this would enable robust comparisons between most subgroups, and accepted that we would only be able to identify large differences between subgroups that contained very few people e.g. those in the highest income category or those reporting a religion other than Christianity or Islam.

We reviewed the number of people who had been recruited on a weekly basis and stopped data collection on the day that the sample exceeded 3,850.

### Statistical analysis

We used logistic regression to calculate the adjusted odds of non-attendance for each sociodemographic subgroup. Our statistical approach is outlined below:

1. Perform simple logistic regression with attendance as the outcome. Separately add each sociodemographic domain as an exposure. (Unadjusted model)
2. Adjust each model for age and gender. (Minimally adjusted model)
3. Adjust each model for all other sociodemographic variables. (Fully adjusted model)
4. Test an interaction between each sociodemographic variable and age category (Effect modification by age)
5. Test an interaction between each sociodemographic variable and gender (Effect modification by gender)

### Post-hoc sensitivity analyses

To quantify the impact of intersectionality,^25,26^ we estimated the probability of attendance for people with different combinations of sociodemographic characteristics that were found to be the strongest predictors for poor access.

After completing our analysis, our Kenyan Ministry of Health collaborators sensibly hypothesized that severity of eye condition could explain differences in attendance by age and other sociodemographic domains, reasoning that those with painful or severe conditions would be more likely to seek care than those with mild or painless conditions. Data on eye conditions had already been collected during screening. We categorised these diagnostic codes into five categories that grouped conditions based on their likely acuity and impact (below). Then we re-ran the regression models with and without this new eye condition data.

- Normal vision
- Loss of vision (visual acuity <6/12 vision in either eye)
- Chronic problem: Growth on eyeball, Lump on lids, White pupil, Strabismus
- Acute problem: Conjunctivitis, Redness, Redness with discharge, Red and watery itchy eye
- Urgent problem: Eye injury, Pain, Whole eyeball swollen

### Bias

To reduce the risk of selection bias the sociodemographic questions were asked of every consecutive person who was referred until we had collected data from at least 3,850 people. We developed a robust set of questions to minimise the risk of recall bias, grounded in the literature and tailored to the local context by a group of experts and community representatives. We delivered standardised training to the data collectors in order to minimise the risk of measurement bias. We also performed unannounced observations of screeners to check that the questions were being asked as intended. We found no issues.

### Ethics

This study was approved by LSHTM and KEMRI ethics committee and the National Commission for Science, Technology & Innovation. Written informed consent was obtained from every participant.

### Findings

Between April and July 2023, 136,912 people aged >18 years old were screened in Meru county and 32,835 people were found to have an eye problem that required referral to a local treatment outreach clinic (24.0%). We gathered and analysed data from the first 4,240 of these referred people who consented to provide their sociodemographic information. As several hundred people were screened every week, our final sample exceeded 3,850.

Of these 4,240 people, just under half were able to access their appointment (46.0%). In our fully adjusted model, we found very strong evidence (p<0.001) of an association between three variables and access: gender, with men found to be less likely to access care than women; age, with younger people less likely to access care than older people; and occupation, where those in skilled/unskilled manual labour and sales & services occupations had the lowest access. Younger people had the worst access overall, with only 32% of those aged 18-44 years being checked-in at clinics compared to 54% of those aged ≥45 years old.

Three other variables showed some weaker evidence of an association with the outcome; education (p=0.03), marital status (p=0.03), and vehicle ownership (p=0.03). (Table 2)

**Table 2:**
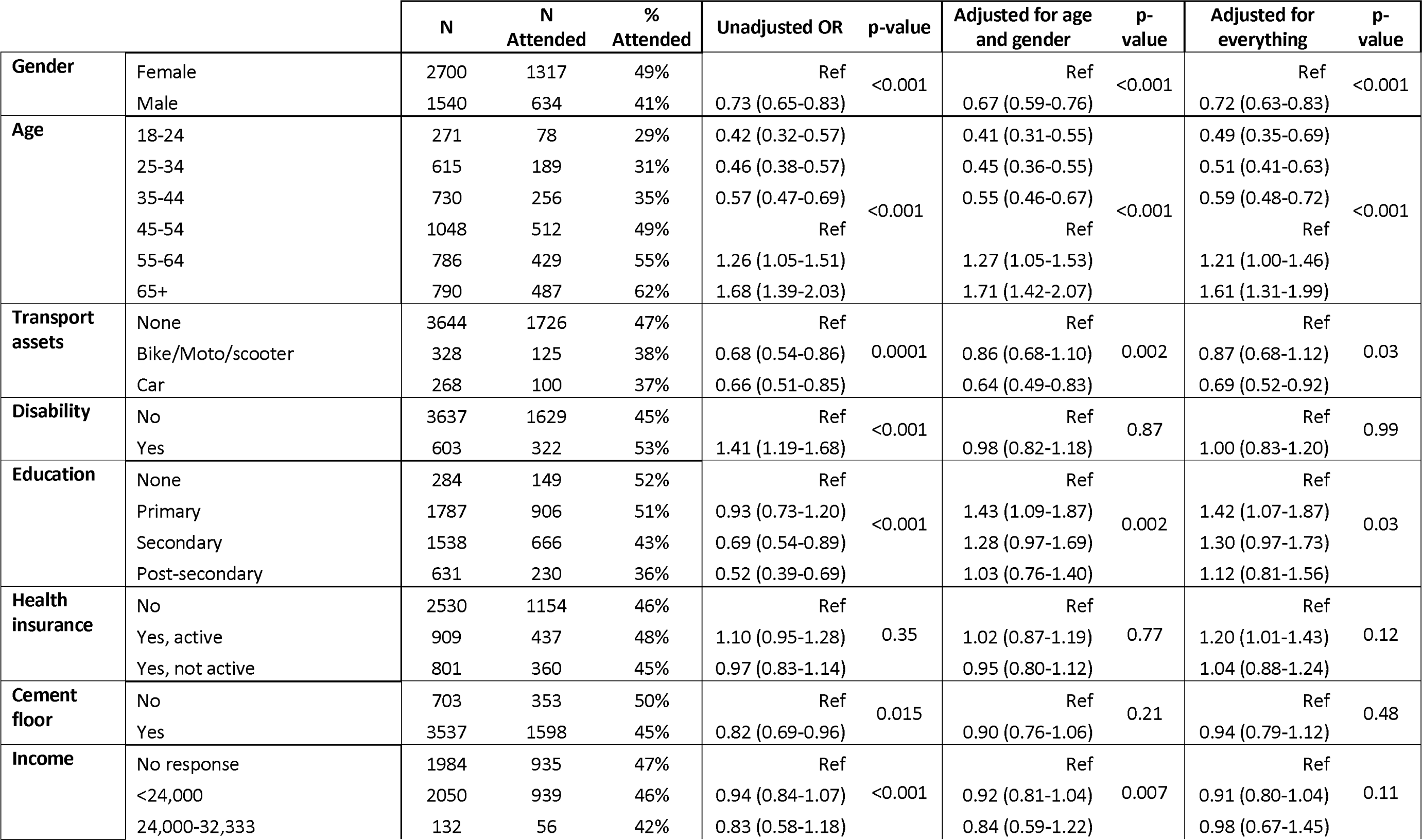
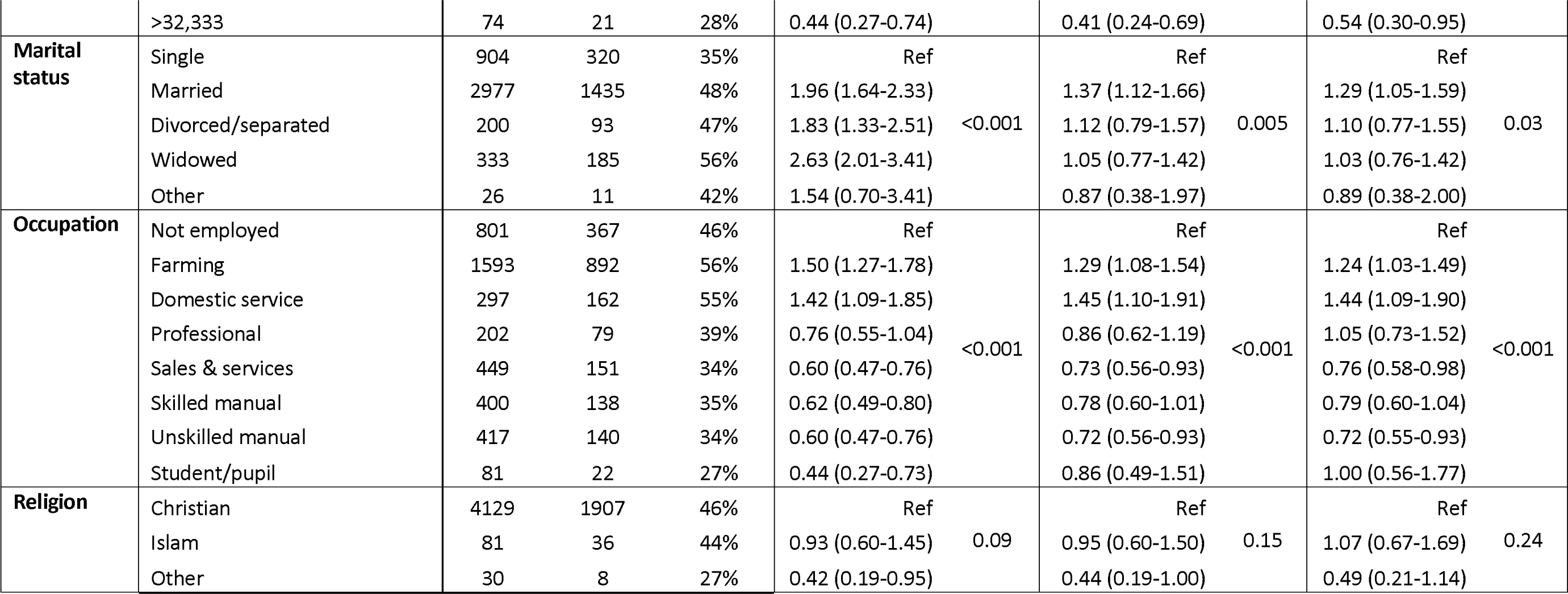
Attendance by sociodemographic group.

Figures 1 and 2 plot the adjusted odds ratios of attendance for the demographic and economic factors.

**Figure 1:**
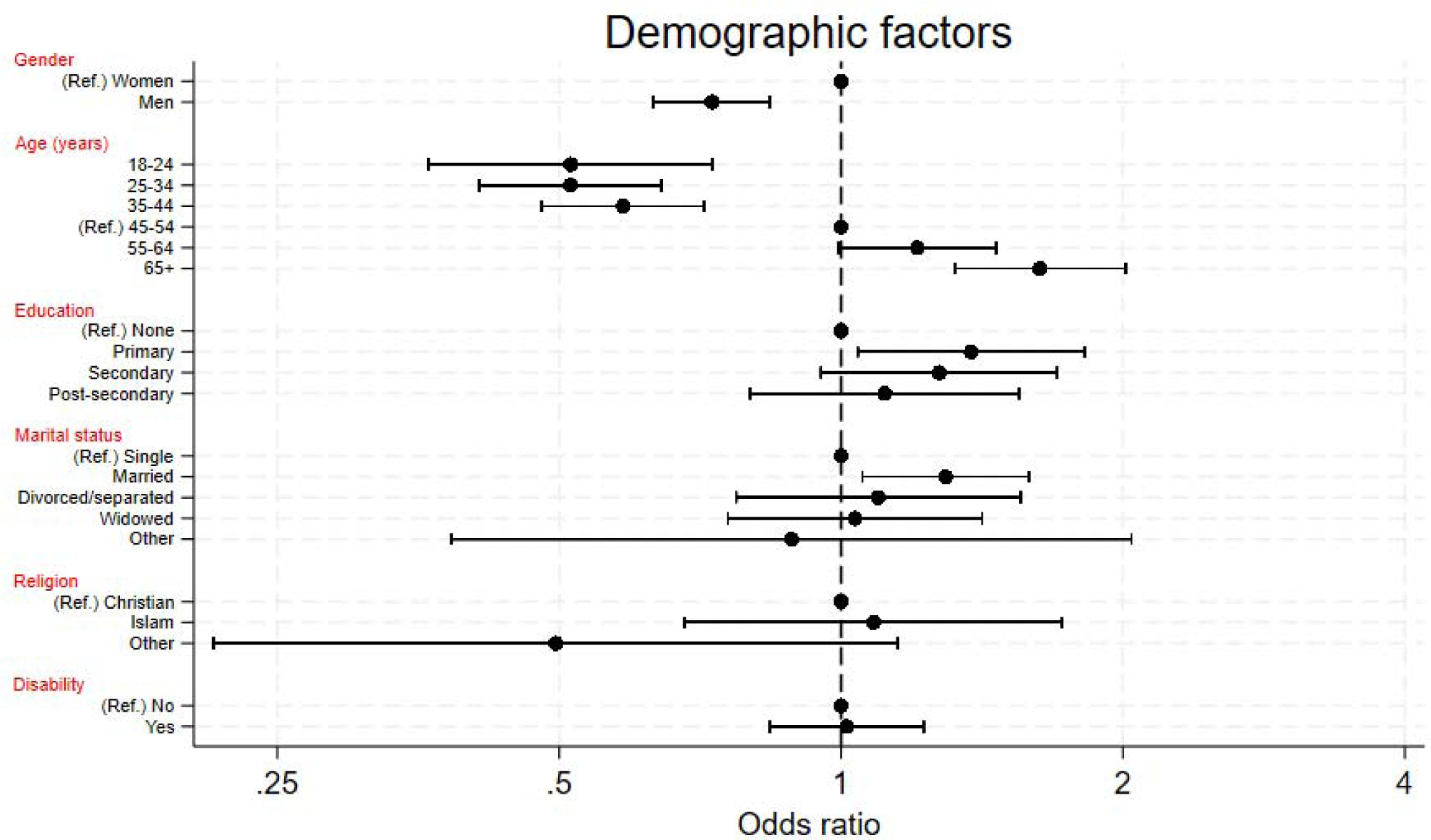
Plot of fully adjusted odds ratios of attendance according to demographic factors. Ref. = Reference group, disability = yes means the participant responded that they had difficulty with at least one of hearing, walking, climbing steps or communicating

**Figure 2:**
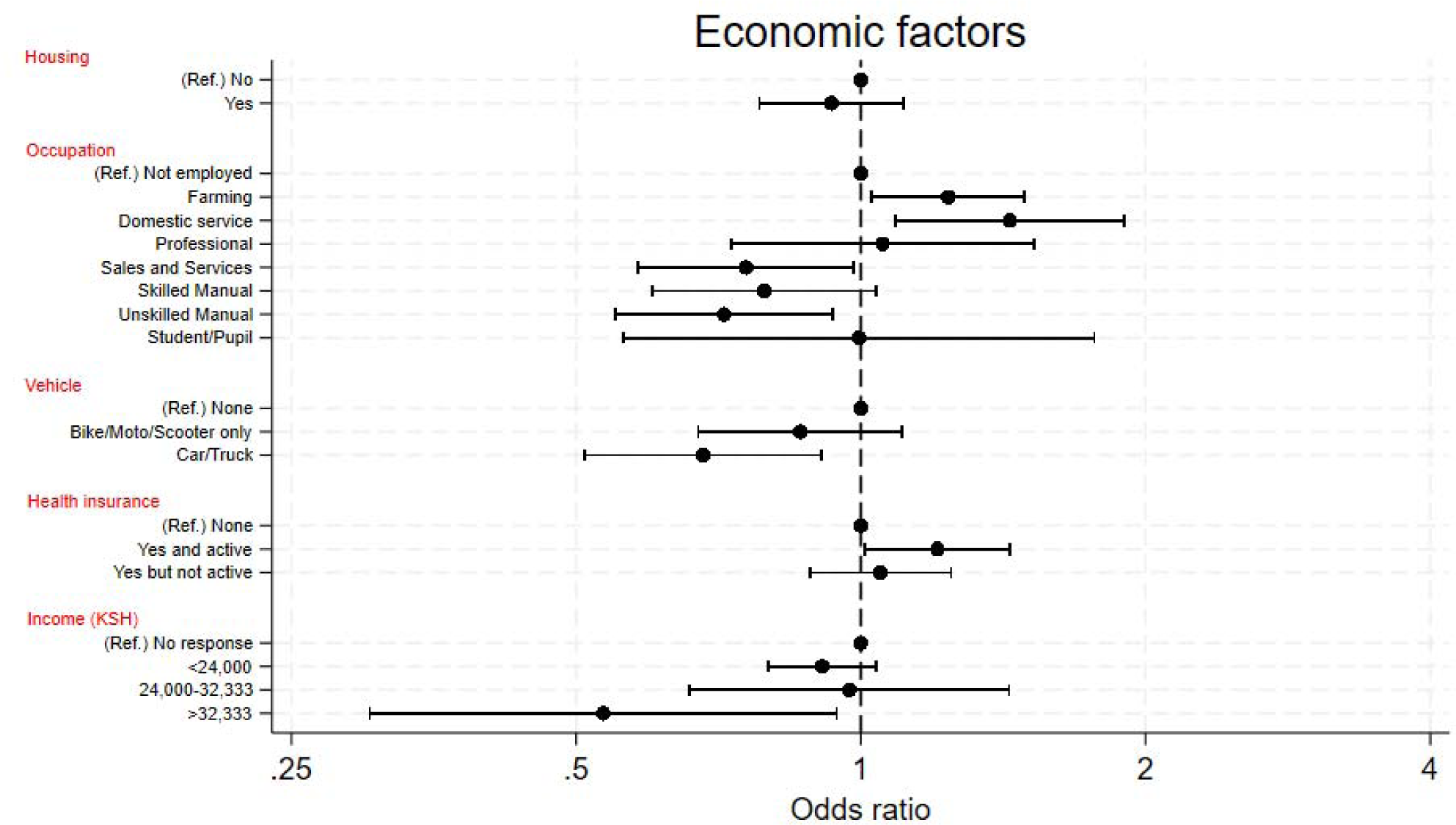
Plot of fully adjusted odds ratios of attendance according to economic factors. Ref. = Reference group

We tested for effect modification and identified some weak evidence (p=0.05) of an interaction between age and gender, suggesting that the difference in attendance between men and women is greater at younger ages than in older (Figure 3 and Supplementary Table 5).

**Figure 3:**
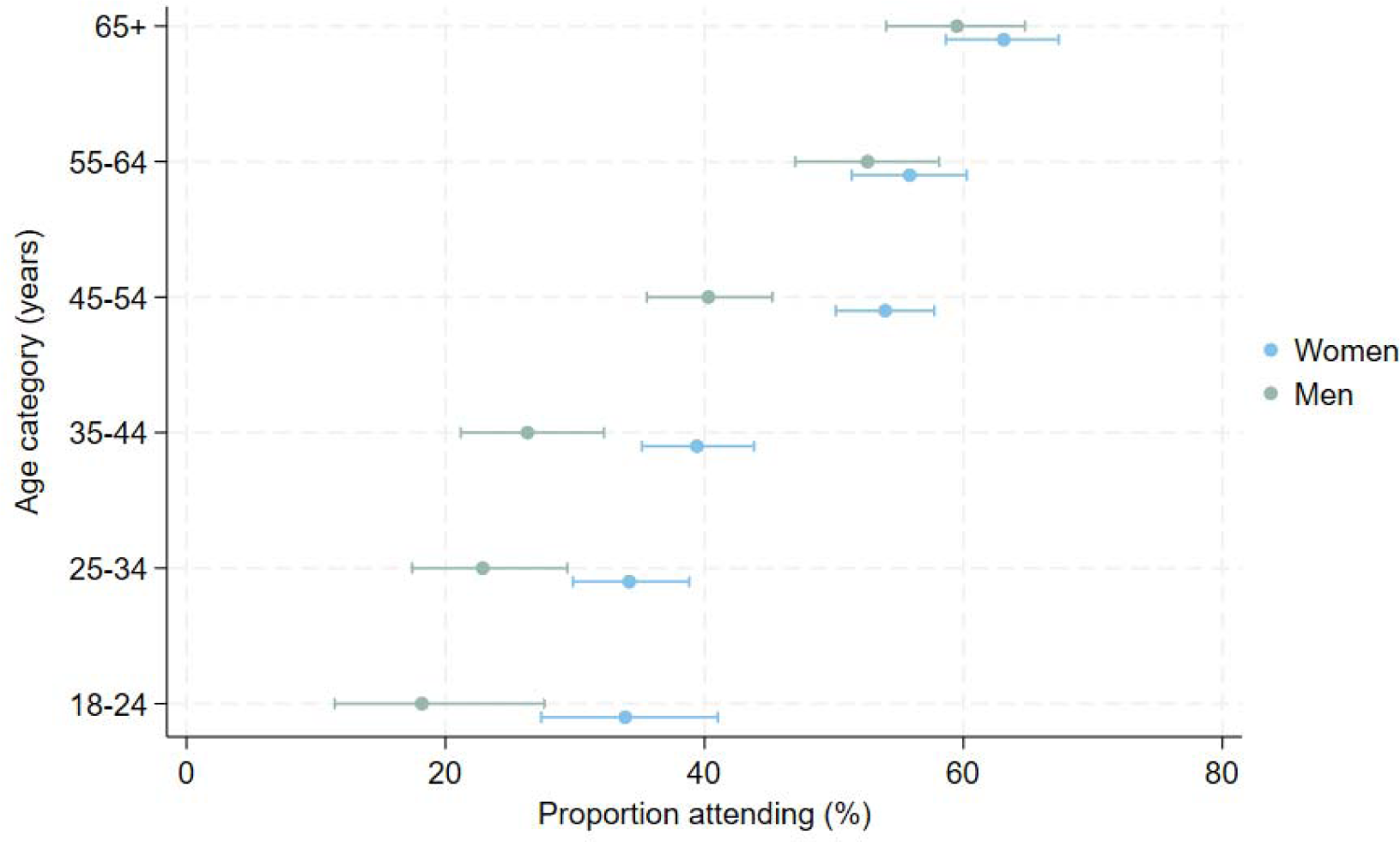
Clinic attendance within each age and gender group.

### Sensitivity analyses

To quantify the impact of intersectionality, we estimated the probability of attendance for people with different combinations of age, gender (including the interaction between age and gender), and occupation – the three strongest predictors of access. Age and gender were already categorical variables. For simplicity, we dichotomised occupation into a binary variable, grouping together the three categories of occupation that had the lowest attendance (skilled/unskilled manual and sales & services).

We found that the expected lowest attending group is 18-24-year-old males who work in sales/service/manual jobs, where we estimate that only 14% of people with these three characteristics would be able to access care (95% CI: 8-22%). The highest estimated access rate was 64%, found among females aged 65+ not working in those occupations (95% CI: 59-68%).

In our second sensitivity analysis we adjusted for severity of eye condition. We found that eye condition did not affect the effect estimates, suggesting that this variable was not driving greater attendance in older people. Supplementary table 6 presents the full results.

## Discussion

The growing emphasis on extending Universal Health Coverage and ‘leaving no-one behind’ means that programme managers around the world are increasingly being expected to identify populations that face unique barriers to care. Aligning with findings from previous research in Kenya,^27^ we found that less than half of all people who screened positive in Meru’s VIP project were able to access care. This resonates with a 2018 systematic review that found that 43% of all African outpatient appointments are not attended, with younger adults and those from lower socioeconomic groups being the least likely to attend.^28^

We found that younger men working in sales, services, or manual jobs were the least likely to attend. This stands in stark contrast to existing research on access to eye services which has shown older age, female gender, and widowhood to be the strongest predictors of poor access.^7,10^ However, these studies focused on cataract care which affects people later on in life, whereas the VIP programme manages all eye conditions in all ages.

Given that Kenya ranks 110^th^ out of 144 countries in the UN’s gender equality ranking,^29^ we were surprised that men were 30% less likely to attend than women in the fully adjusted model. However, this is not an unusual finding: despite having greater power, privileges, and opportunities than women in virtually all societies, men almost universally experience higher rates of poor health, lower rates of health care access, and lower overall life expectancy.^30,31^ Differences in healthcare-seeking behaviour are thought to drive much of the gender gap in access rates, related to differences in perception of risk and pervasive social ideals of masculinity.^32^ Whilst younger men were the least likely to attend in Meru, younger women were less likely to attend than older women, suggesting that youth is an independent predictor. Overall, age was by far the strongest predictor, with the youngest cohort (18-24y) three times less likely to have been checked-in than the oldest (65+), even after adjusting for occupation and severity of eye condition.

We hypothesise that younger adults may be more likely to be ‘hustling’ than older people – i.e. working in informal jobs with no fixed salary or paid sick leave, and therefore facing higher financial opportunity costs when taking time out to attend a clinic. The fact that people working in (often informal) sales, services, and manual labour were also less likely to attend than those working in other areas seems to corroborate this hypothesis.

To a lesser extent, car/truck ownership and high level of income were also associated with poor access. We hypothesise that this is because richer people who are told they have an eye problem at screening may be seeking private care rather than attending the VIP clinics. We plan to conduct a further set of interviews with people from this group to explore this issue further.

Our study had a number of limitations. We did not include questions on religion, tribe/ethnicity, or sexuality due to concerns about cultural sensitivities, but these are all important markers of potential access challenges.^17,18^ With a larger sample we would have been able to detect smaller differences between groups, however it would have taken longer to conduct the study and the embedded nature of this research comes with pressure to deliver rapid and timely findings. Finally, we have not yet validated our sociodemographic questions. This work is currently underway, however the process of selecting the items and response options was based on extensive literature review and wide stakeholder engagement to ensure that we were using previously-validated questions with strong external validity.

## Conclusion

Less than half of those referred to local eye clinics received treatment. We found evidence of large sociodemographic inequalities, with younger people, males, and those working in sales, services, and manual jobs facing the highest barriers. Overall, age was the strongest predictor. Future work should focus on exploring the specific barriers faced by younger adults and their ideas for how services could be modified to improve access to essential eye care.

## Data Availability

Patient-level data will be pseudo-anonymised removing names and any other key identifiers before it is shared. Only the least amount of data will be shared, and where possible it will be fully anonymised and aggregated. All published findings will be at anonymous aggregate subpopulation level. In line with the UK concordat on open research data (2016), anonymised data from this trial will be made available to bona fide research groups (evidenced via CVs and the involvement of a qualified statistician)

## Appendix

**Supplementary Table 1:**
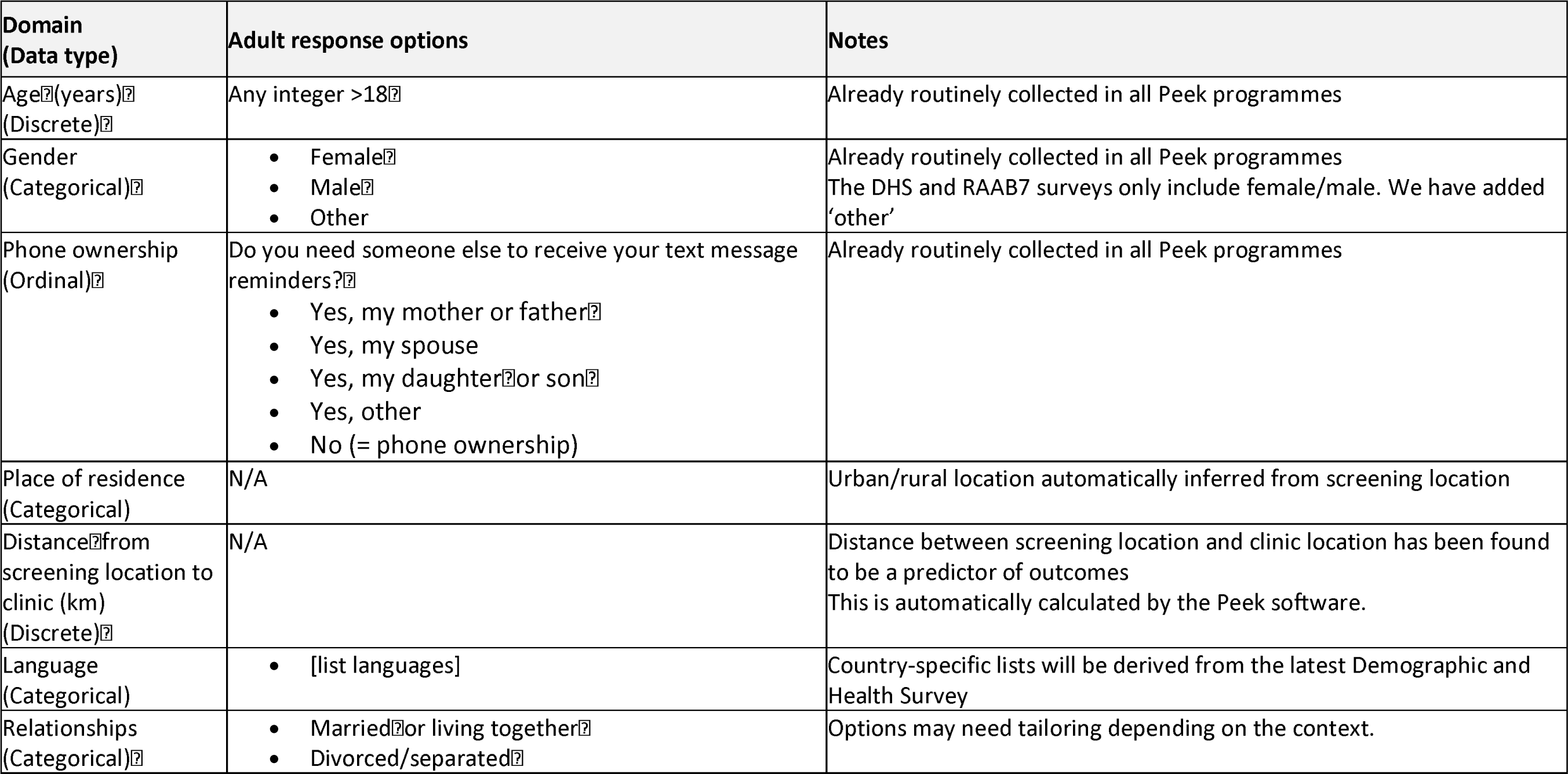
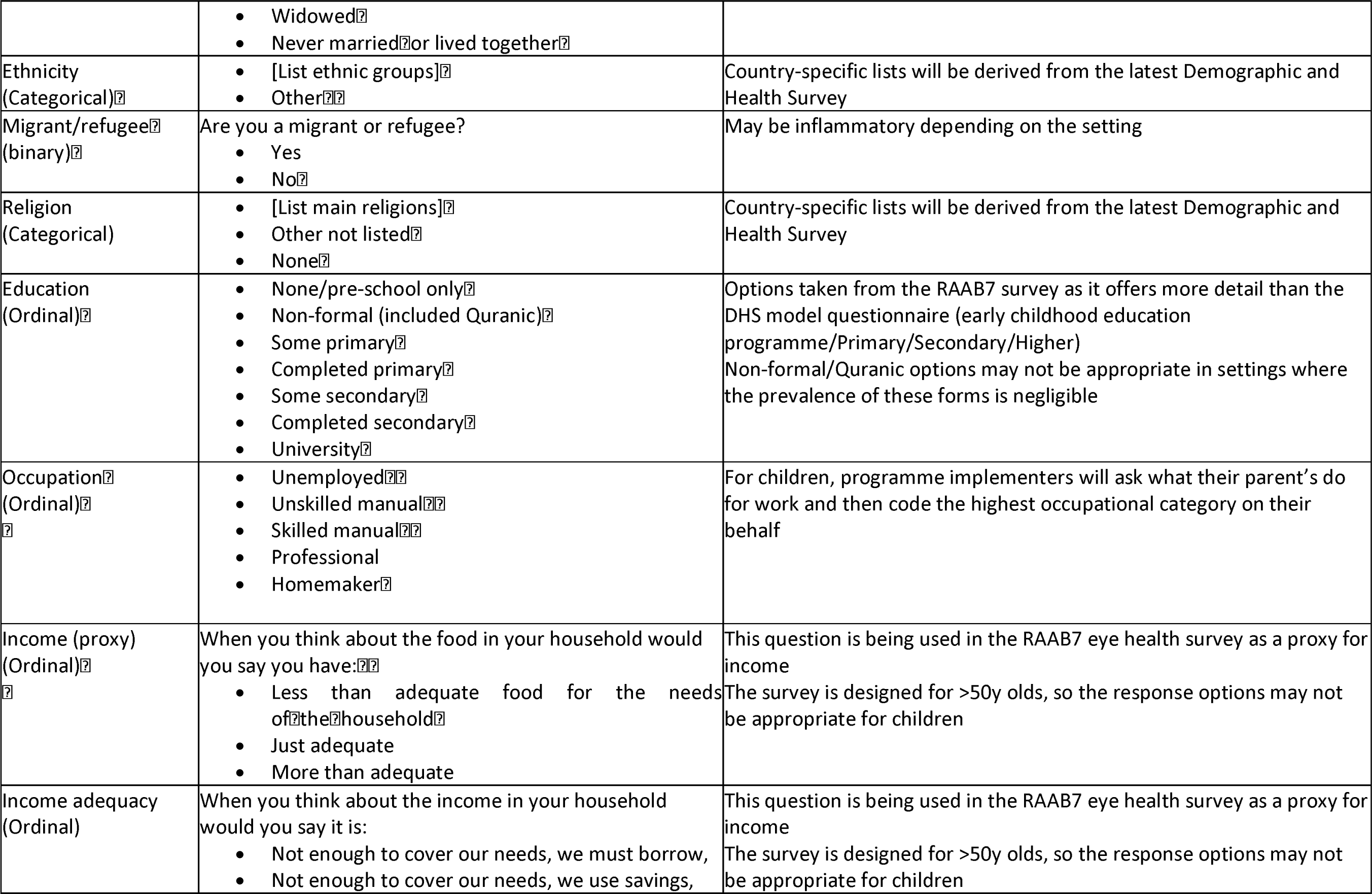
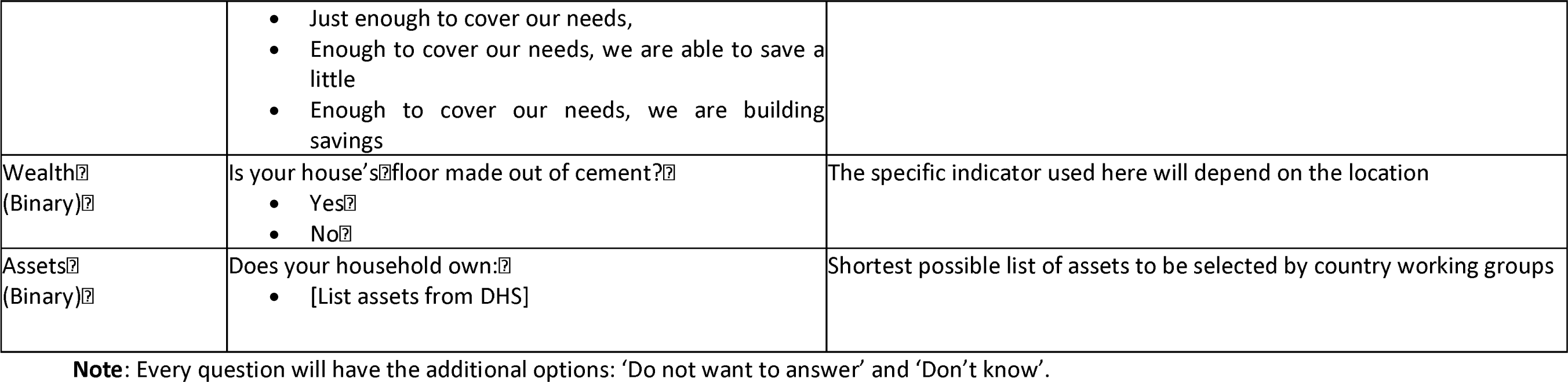
Sociodemographic variables from the first multi-stakeholder workshop.

**Supplementary Table 2:** Sociodemographic variables from the second multi-stakeholder workshop.

| Domain | Adult response options | Notes |
| --- | --- | --- |
| Age | Any integer >18 | Already routinely gathered |
| Gender | •Female<br>•Male<br>•Other | Already routinely gathered |
| Phone ownership | Do you need someone else to receive your text? message reminders?<br>• Mother or father<br>• Spouse<br>• Daughter or son<br>• Other<br>• No (=phone ownership) | Already routinely gathered |
| Place of residence | N/A | Urban/rural automatically inferred |
| Distance to clinic | N/A | Automatically calculated by Peek |
| Language | What language do you speak most often at home?<br>•English<br>•Swahili<br>•Borana<br>•Embu<br>•Kalenjin<br>•Kamba<br>•Kikuyu<br>•Kisii<br>•Luhya<br>•Maragoli<br>•Luo | Workshop participants felt that it would be inflammatory to ask about tribe/ethnicity. Language will be used as a Proxy |
|  | •Maasai<br>•Meru<br>•Mijikenda<br>•Pokot<br>•Somali<br>•Turkana<br>•Other |  |
| Relationships | •Never married<br>•Married ☐<br>•Living together<br>•Single<br>•Divorced/separated<br>•Widowed ☐ |  |
| Migrant status | Were you born in Kenya?<br>•Yes<br>•No<br>•Don't want to answer | This question may be redundant. Kenya is currently home to 500,000 refugees, however, they mainly live in camps and this information will already be collected under 'Place of residence'. Outside of Nairobi, the migrant population that does not live in camps is negligible. |
| Religion | What is your religion?<br>•Roman Catholic<br>•Protestant/other Christian<br>•Islam<br>•Other<br>•No religion | Responses taken from the 2014 DHS |
| Education | What is you highest level of completed schooling?<br>•No education<br>•Some primary<br>•Primary complete<br>•Some secondary<br>•Secondary complete<br>•More than secondary | Adult responses aligned with the 2014 DHS |
| Occupation | What is your occupation?<br>•Unemployed<br>•Agriculture<br>•Unskilled manual<br>•Skilled manual<br>•Sales and services<br>•Clerical<br>•Professional/technical/managerial<br>•Homemaker | Interviewer to categorise and code the highest |
| Food adequacy | When you think about the food in your household would you say you have:<br>• Less than adequate food for the needs of the household<br>• Just adequate<br>• More than adequate | Question taken from RAAB7<br>– may remove due to poor face validity |
| Income adequacy | When you think about the income in your household would you say it is:<br>• Not enough to cover our needs, we must borrow,<br>• Not enough to cover our needs, we use savings,<br>• Just enough to cover our needs,<br>• Enough to cover our needs, we are able to save a little<br>• Enough to cover our needs, we are building savings | From RAAB7, but poor face validity. |
| Housing | Is your house's floor made of earth, sand, or dung?<br>• Yes<br>• No<br>Do you have water piped into your own house or yard?<br>• Yes<br>• No<br>Does your household have electricity?<br>• Yes<br>• No<br>What kind of toilet does your household you use?<br>• Own toilet/latrine<br>• Shared toilet/latrine<br>• None (bush/field) | All options taken from the 2014 DHS |
| Assets | Do you own a smartphone?<br>•Yes<br>•No<br>Does your household own a:<br>•Bicycle<br>•Motorcycle/scooter<br>•Car or truck<br>Do you own your dwelling?<br>•Yes<br>•No |  |

**Supplementary Table 3:** Sociodemographic variables from the third multi-stakeholder workshop.

| Domain | Adult response options | Child response options | Notes |
| --- | --- | --- | --- |
| Age | Any integer >18 | Any integer 5 - 17 | Already routinely gathered |
| Gender | •Female<br>•Male<br>•Other | •Female<br>•Male<br>•Other | Already routinely gathered |
| Phone ownership | Do you need someone else to receive your text? message reminders?<br>• Mother or father<br>• Spouse<br>• Daughter or son<br>• Other<br>• No (=phone ownership) | Provided contact number:<br>• Mother or father<br>• Guardian<br>• Teacher<br>• Other | Already routinely gathered |
| Place of residence | N/A | N/A | Urban/rural automatically inferred |
| Distance to clinic | N/A | N/A | Automatically calculated by Peek |
| Language | What language do you speak most often at home?<br>•English<br>•Swahili<br>•Borana<br>•Embu<br>•Kalenjin<br>•Kamba<br>•Kikuyu<br>•Kisii<br>•Luhya<br>•Maragoli<br>•Luo | What language do you speak most often at home?<br>•English<br>•Swahili<br>•Borana<br>•Embu<br>•Kalenjin<br>•Kamba<br>•Kikuyu<br>•Kisii<br>•Luhya<br>•Maragoli | Used instead of ethnicity |
|  | •Maasai<br>•Meru<br>•Mijikenda<br>•Pokot<br>•Somali<br>•Turkana<br>•Other | •Luo<br>•Maasai<br>•Meru<br>•Mijikenda<br>•Pokot<br>•Somali<br>•Turkana<br>•Other |  |
| Relationships | •Never married<br>•Married?<br>•Living together<br>•Single<br>•Divorced/separated<br>•Widowed? | *Do you live with:*<br>• Both parents?<br>• Just one parent?<br>• Another relative<br>• Guardian?(non-relative)<br>• Orphanage |  |
| Religion | What is your religion?<br>•Roman Catholic<br>•Protestant/other Christian<br>•Islam<br>•Other<br>•No religion | What is your religion?<br>•Roman Catholic<br>•Protestant/other Christian<br>•Islam<br>•Other<br>•No religion | Responses taken from the 2014 DHS |
| Education | What is your highest level of completed schooling?<br>•No education<br>•Some primary<br>•Primary complete<br>•Some secondary<br>•Secondary complete<br>•More than secondary | N/A | Adult responses aligned with the 2014 DHS |
| Occupation | What is your occupation?<br>•Unemployed<br>•Agriculture<br>•Unskilled manual<br>•Skilled manual<br>•Sales and services<br>•Clerical<br>•Professional/technical/managerial<br>•Homemaker | What are your parents' jobs?<br>•No parents<br>•Unemployed<br>•Agriculture<br>•Unskilled manual<br>•Skilled manual<br>•Sales and services<br>•Clerical<br>•Professional/technical/managerial<br>•Homemaker | Interviewer to categorise and code the highest |
| Income adequacy | When you think about the income in your household would you say it is:<br>• Not enough to cover our needs, we must borrow,<br>• Not enough to cover our needs, we use savings,<br>• Just enough to cover our needs,<br>• Enough to cover our needs, we are able to save a little<br>• Enough to cover our needs, we are building savings | N/A | From RAAB7, but poor face validity. |
| Housing | Is your house's floor made of earth, sand, or dung?<br>•Yes<br>•No<br>Do you have water piped into your own house or yard?<br>•Yes<br>•No<br>Does your household have electricity?<br>•Yes<br>•No<br>What kind of toilet does your household you use?<br>•Own toilet/latrine<br>•Shared toilet/latrine<br>•None (bush/field) | Is your house's floor made of earth, sand, or dung?<br>•Yes<br>•No<br>Do you have water piped into your own house or yard?<br>•Yes<br>•No<br>Does your household have electricity?<br>•Yes<br>•No<br>What kind of toilet does your household you use?<br>•Own toilet/latrine<br>•Shared toilet/latrine<br>•None (bush/field) | All options taken from the 2014 DHS |
| Assets | Do you own a smartphone?<br>•Yes<br>•No<br>Does your household own a:<br>•Bicycle<br>•Motorcycle/scooter<br>•Car or truck<br>Do you own your dwelling?<br>•Yes<br>•No | Does your household own a smartphone?<br>•Yes<br>•No<br>Does your household own a:<br>•Bicycle<br>•Motorcycle/scooter<br>•Car or truck |  |

**Supplementary Table 4:**
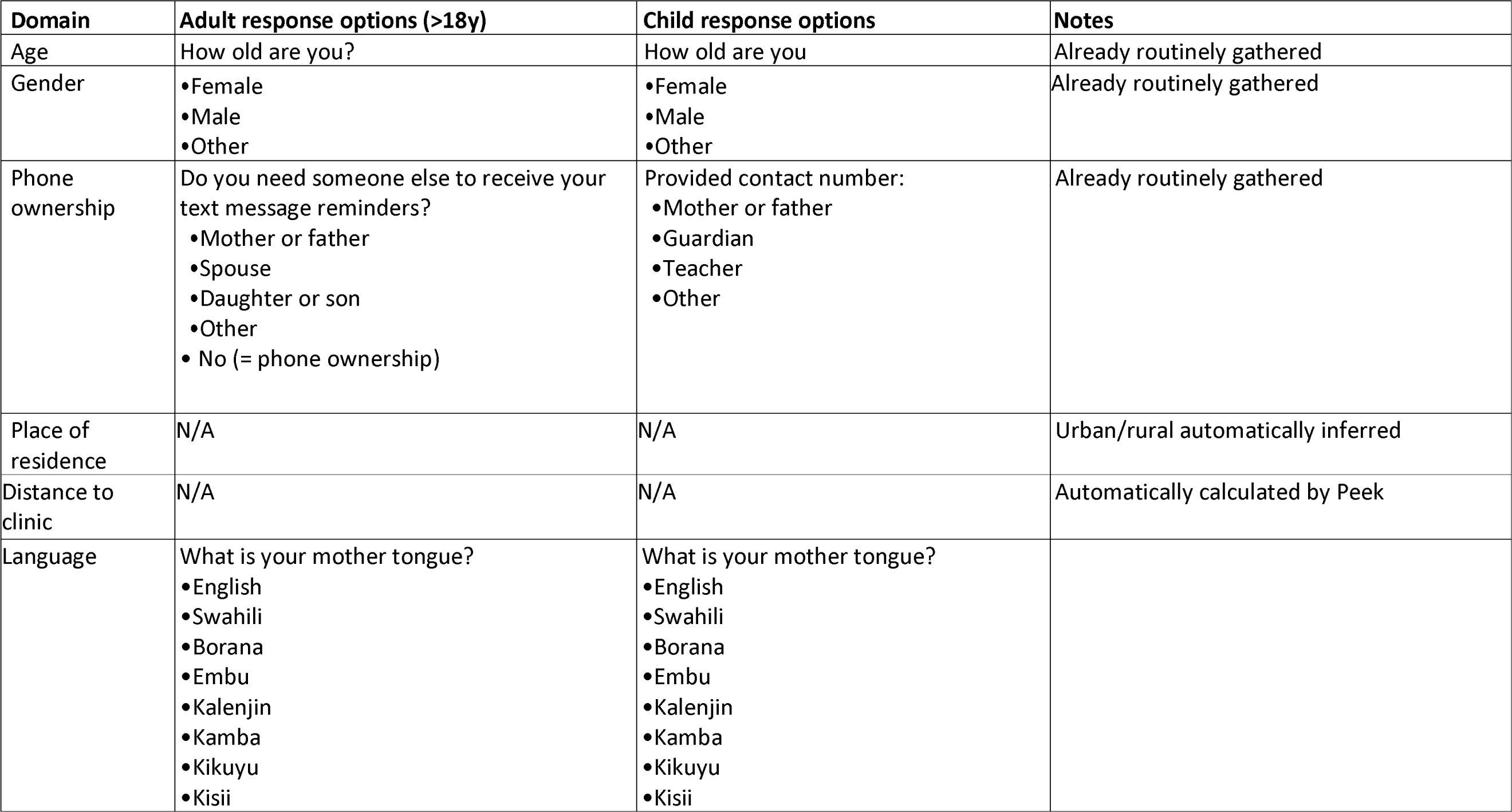
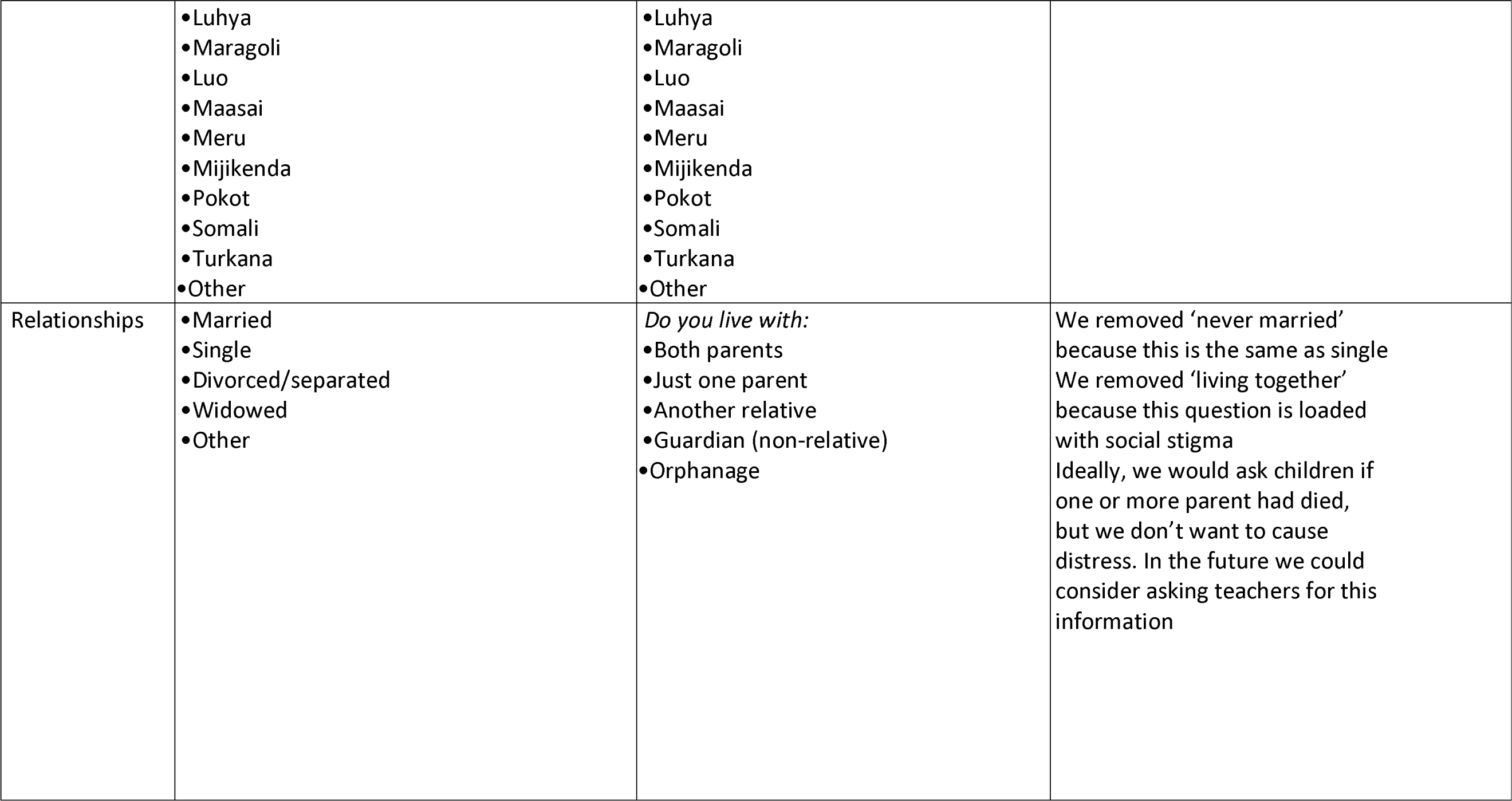
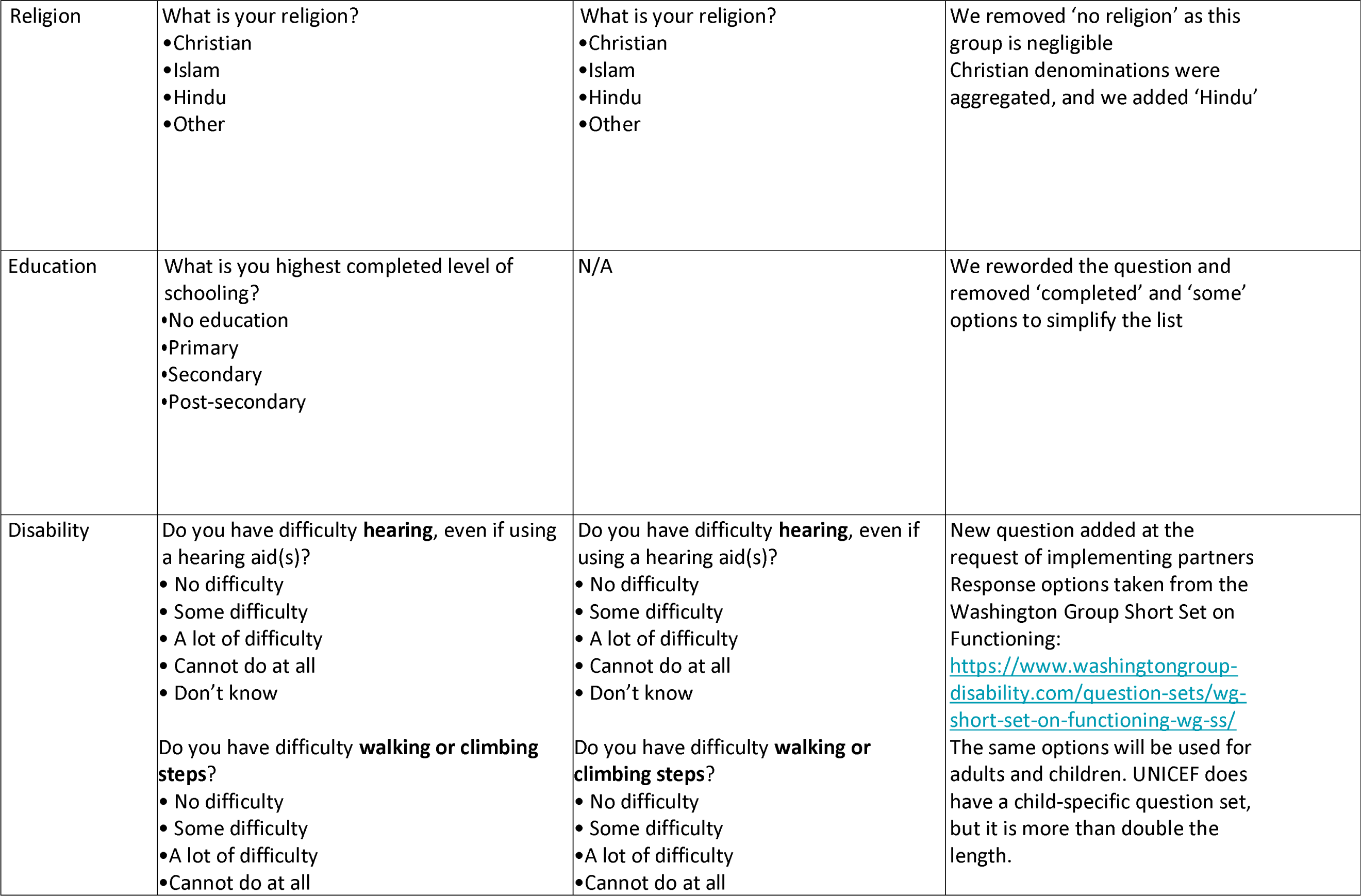
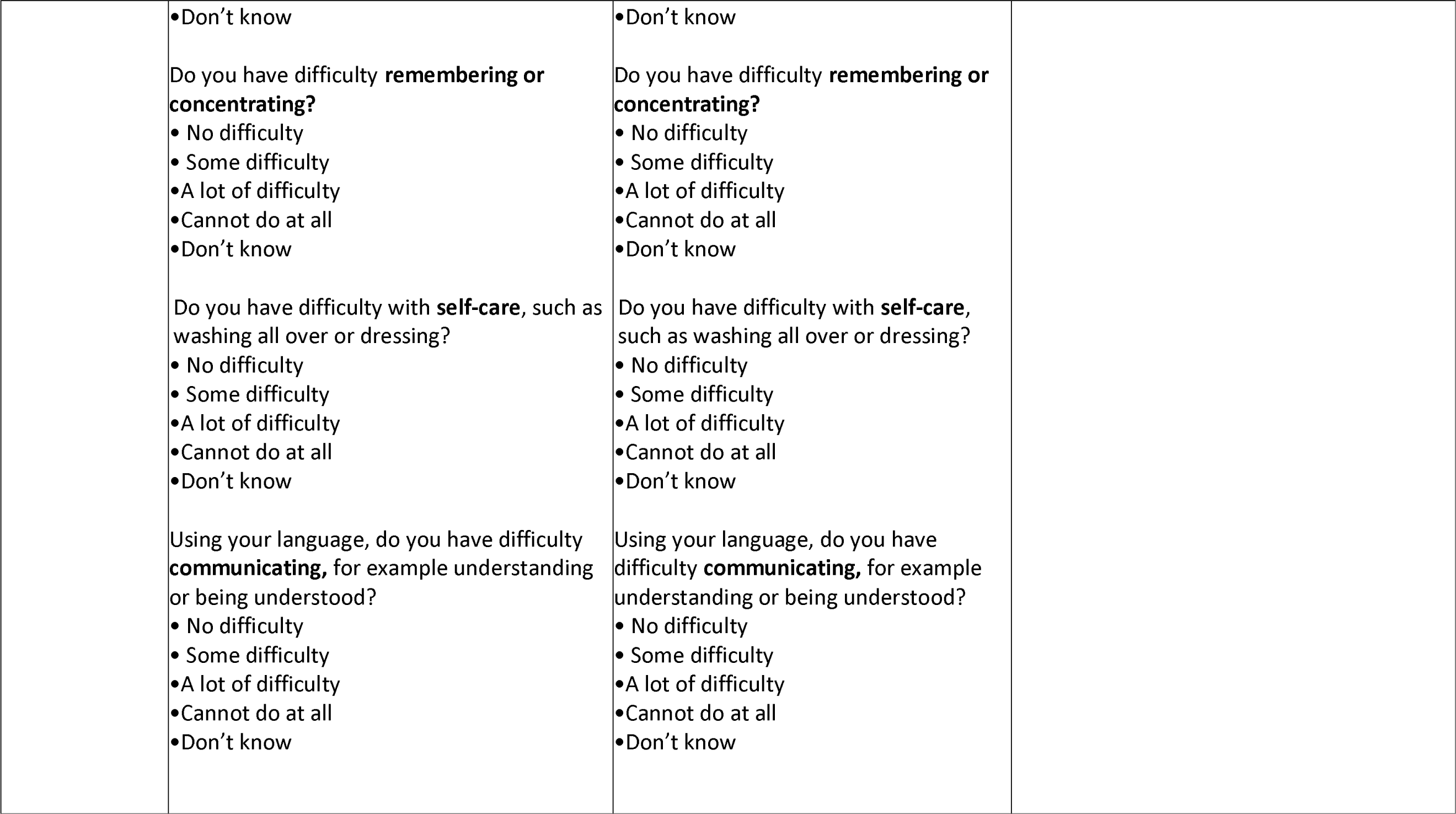
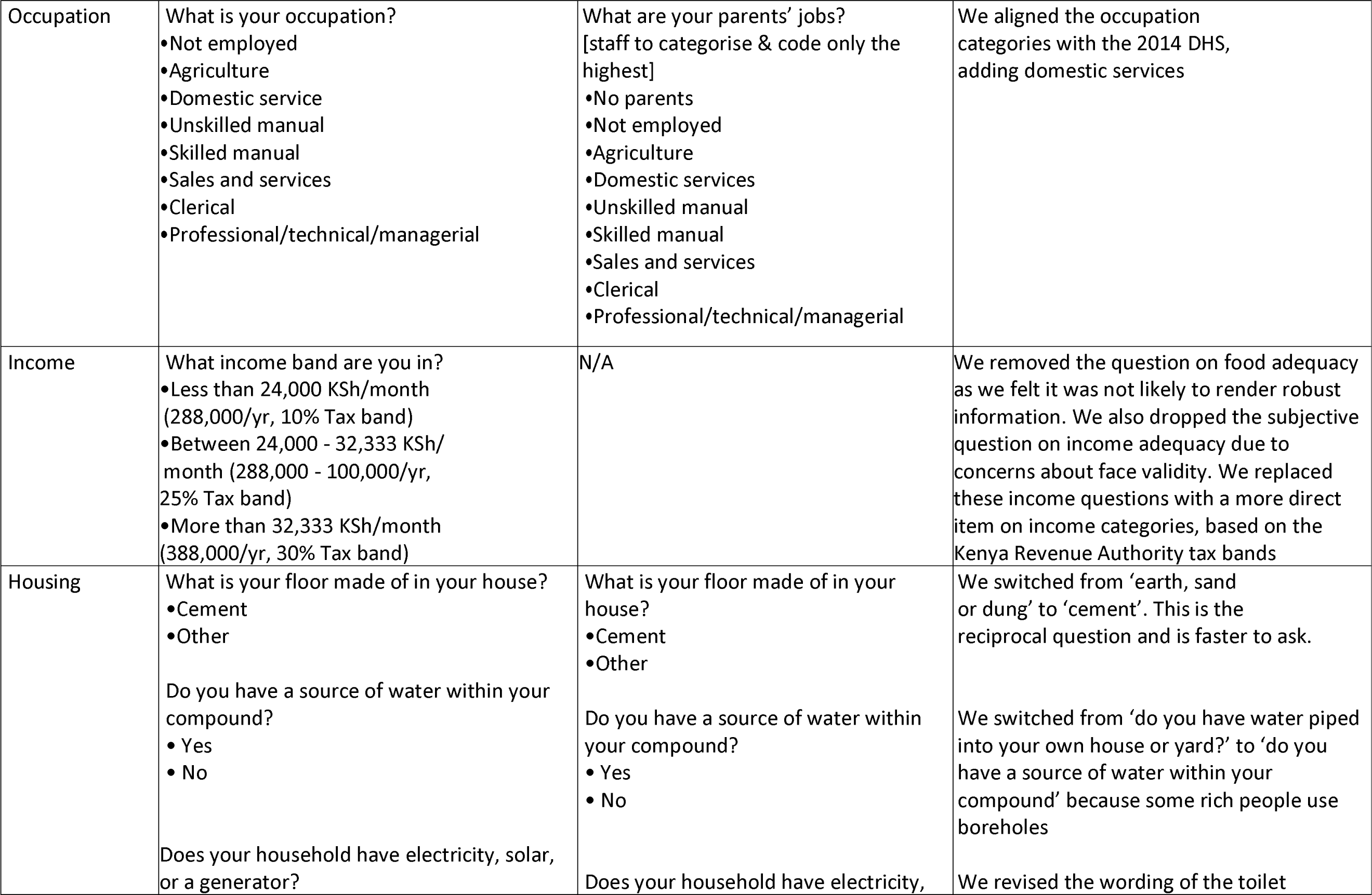
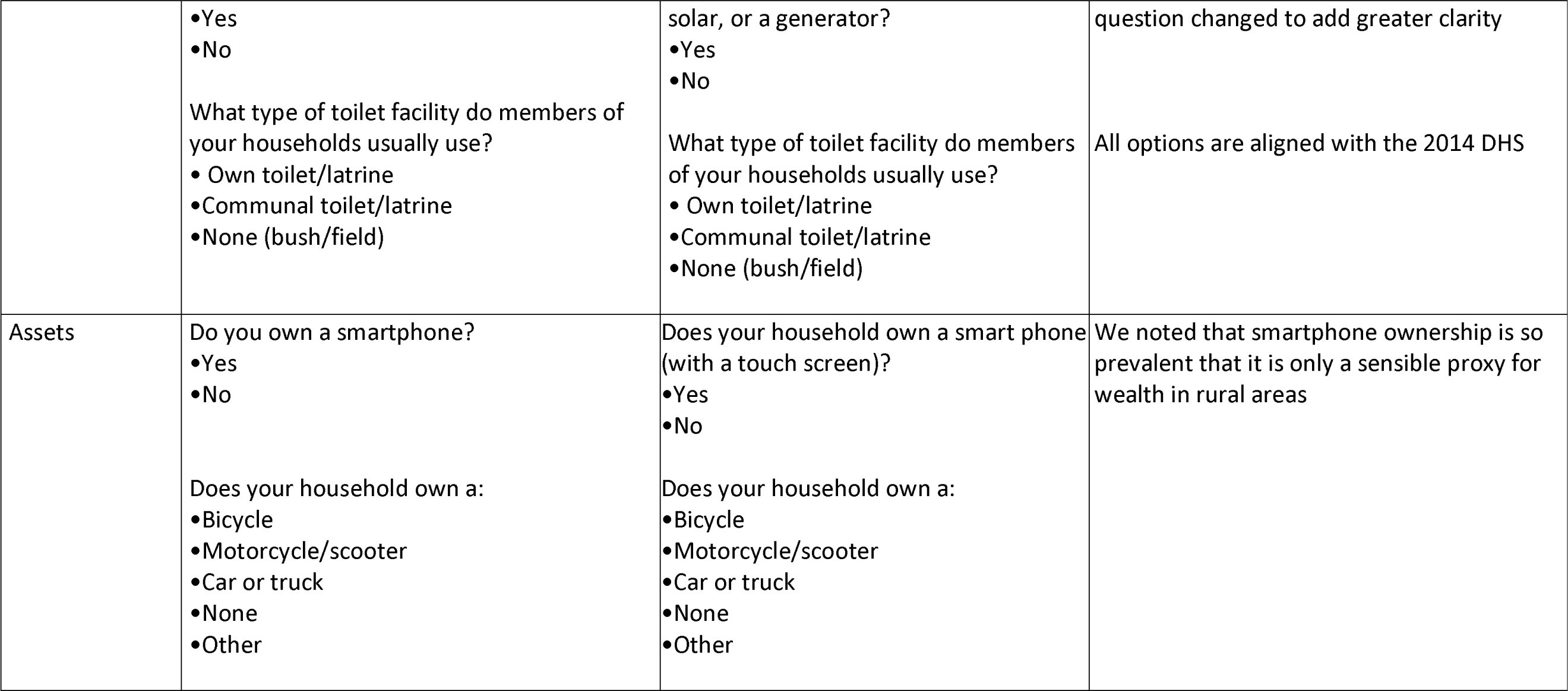
Sociodemographic variables from the fourth multi-stakeholder workshop.

**Supplementary table 5:**
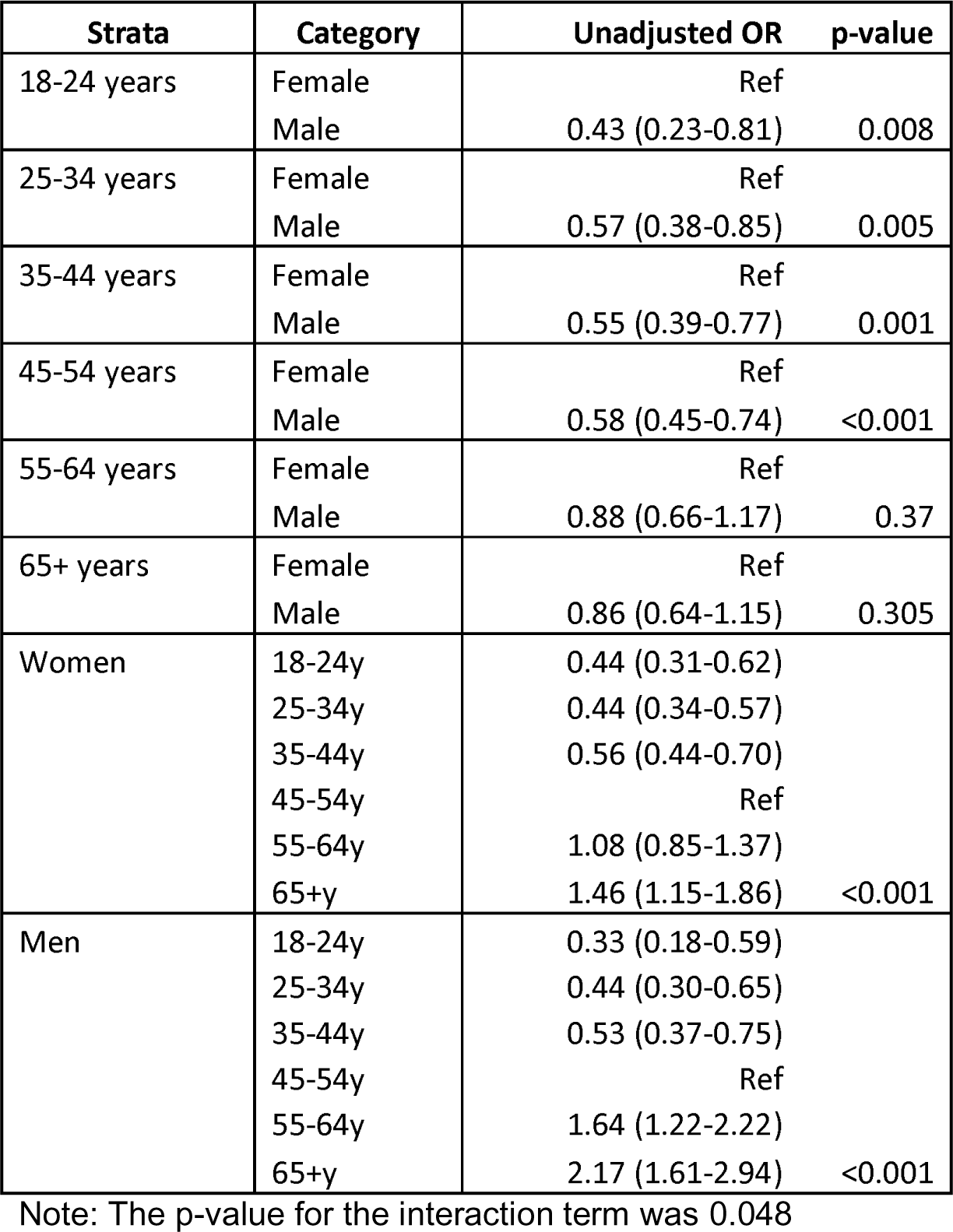
stratum specific effect estimates of association between attendance and age and gender.

**Supplementary table 6:** Regression with additional adjustment for eye condition.

|  |  | N | N Attended | % Attended | Unadjusted | p | Adjusted for age & sex | p | Adjusted for everything | p | Additionally adjusted for eye condition | p |
| --- | --- | --- | --- | --- | --- | --- | --- | --- | --- | --- | --- | --- |
| Gender | F | 2700 | 1317 | 49% | Ref | <0.001 | Ref | <0.001 |  |  | Ref | <0.001 |
|  | M | 1540 | 634 | 41% | 0.73 (0.65-0.83) |  | 0.67 (0.59-0.76) |  | 0.72 (0.63-0.83) | <0.001 | 0.71 (0.62-0.83) |  |
| Age cat | 18-24 | 271 | 78 | 29% | 0.42 (0.32-0.57) |  | 0.41 (0.31-0.55) |  | 0.49 (0.35-0.69) |  | 0.54 (0.38-0.77) | <0.001 |
|  | 25-34 | 615 | 189 | 31% | 0.46 (0.38-0.57) |  | 0.45 (0.36-0.55) |  | 0.51 (0.41-0.63) |  | 0.55 (0.43-0.69) |  |
|  | 35-44 | 730 | 256 | 35% | 0.57 (0.47-0.69) |  | 0.55 (0.46-0.67) |  | 0.59 (0.48-0.72) |  | 0.61 (0.49-0.74) |  |
|  | 45-54 | 1048 | 512 | 49% | Ref | <0.001 | Ref | <0.001 | Ref | <0.001 | Ref |  |
|  | 55-64 | 786 | 429 | 55% | 1.26 (1.05-1.51) |  | 1.27 (1.05-1.53) |  | 1.21 (1.00-1.46) |  | 1.20 (0.99-1.45) |  |
|  | 65+ | 790 | 487 | 62% | 1.68 (1.39-2.03) |  | 1.71 (1.42-2.07) |  | 1.61 (1.31-1.99) |  | 1.65 (1.33-2.04) |  |
| Asset | None | 3644 | 1726 | 47% | Ref | 0.0001 | Ref | 0.002 | Ref | 0.03 | Ref | 0.02 |
|  | Bike/Moto/scooter | 328 | 125 | 38% | 0.68 (0.54-0.86) |  | 0.86 (0.68-1.10) |  | 0.87 (0.68-1.12) |  | 0.86 (0.67-1.10) |  |
|  | Car | 268 | 100 | 37% | 0.66 (0.51-0.85) |  | 0.64 (0.49-0.83) |  | 0.69 (0.52-0.92) |  | 0.68 (0.51-0.91) |  |
| Disability | No | 3637 | 1629 | 45% | Ref |  | Ref |  | Ref | 0.99 | Ref | 0.86 |
|  | Yes | 603 | 322 | 53% | 1.41 (1.19-1.68) | <0.001 | 0.98 (0.82-1.18) | 0.87 | 1.00 (0.83-1.20) |  | 1.02 (0.84-1.23) |  |
| Education | None | 284 | 149 | 52% | Ref | <0.001 | Ref | 0.002 | Ref | 0.03 | Ref | 0.05 |
|  | Primary | 1787 | 906 | 51% | 0.93 (0.73-1.20) |  | 1.43 (1.09-1.87) |  | 1.42 (1.07-1.87) |  | 1.37 (1.04-1.82) |  |
|  | Secondary | 1538 | 666 | 43% | 0.69 (0.54-0.89) |  | 1.28 (0.97-1.69) |  | 1.30 (0.97-1.73) |  | 1.27 (0.95-1.70) |  |
|  | Post-secondary | 631 | 230 | 36% | 0.52 (0.39-0.69) |  | 1.03 (0.76-1.40) |  | 1.12 (0.81-1.56) |  | 1.10 (0.79-1.53) |  |
| Health insurance | No | 2530 | 1154 | 46% | Ref | 0.35 | Ref | 0.77 | Ref | 0.12 | Ref | 0.13 |
|  | Yes active | 909 | 437 | 48% | 1.10 (0.95-1.28) |  | 1.02 (0.87-1.19) |  | 1.20 (1.01-1.43) |  | 1.20 (1.01-1.43) |  |
|  | Yes not active | 801 | 360 | 45% | 0.97 (0.83-1.14) |  | 0.95 (0.80-1.12) |  | 1.04 (0.88-1.24) |  | 1.04 (0.88-1.23) |  |
| Housing | No | 703 | 353 | 50% | Ref | 0.015 | Ref | 0.21 | Ref | 0.48 | Ref | 0.43 |
|  | Yes | 3537 | 1598 | 45% | 0.82 (0.69-0.96) |  | 0.90 (0.76-1.06) |  | 0.94 (0.79-1.12) |  | 0.93 (0.78-1.11) |  |
| Income | No response | 1984 | 935 | 47% | Ref | <0.001 | Ref | 0.007 | Ref | 0.11 | Ref | 0.11 |
|  | <24,000 | 2050 | 939 | 46% | 0.94 (0.84-1.07) |  | 0.92 (0.81-1.04) |  | 0.91 (0.80-1.04) |  | 0.91 (0.80-1.04) |  |
|  | 24,000-32,333 | 132 | 56 | 42% | 0.83 (0.58-1.18) |  | 0.84 (0.59-1.22) |  | 0.98 (0.67-1.45) |  | 1.00 (0.67-1.47) |  |
|  | >32,333 | 74 | 21 | 28% | 0.44 (0.27-0.74) |  | 0.41 (0.24-0.69) |  | 0.54 (0.30-0.95) |  | 0.53 (0.30-0.94) |  |
| Marital | Single | 904 | 320 | 35% | Ref | <0.001 | Ref | 0.005 | Ref | 0.03 | Ref | 0.05 |
|  | Married | 2977 | 1435 | 48% | 1.96 (1.64-2.33) |  | 1.37 (1.12-1.66) |  | 1.29 (1.05-1.59) |  | 1.30 (1.06-1.59) |  |
|  | Divorced/separate | 200 | 93 | 47% | 1.83 (1.33-2.51) |  | 1.12 (0.79-1.57) |  | 1.10 (0.77-1.55) |  | 1.11 (0.78-1.58) |  |
|  | Widowed | 333 | 185 | 56% | 2.63 (2.01-3.41) |  | 1.05 (0.77-1.42) |  | 1.03 (0.76-1.42) |  | 1.04 (0.76-1.43) |  |
|  | Other | 26 | 11 | 42% | 1.54 (0.70-3.41) |  | 0.87 (0.38-1.97) |  | 0.89 (0.38-2.00) |  | 0.88 (0.38-2.03) |  |
| Occupation | Not employed | 801 | 367 | 46% | Ref | <0.001 | Ref | <0.001 | Ref | <0.001 | Ref | <0.001 |
|  | Farming | 1593 | 892 | 56% | 1.50 (1.27-1.78) |  | 1.29 (1.08-1.54) |  | 1.24 (1.03-1.49) |  | 1.25 (1.04-1.51) |  |
|  | Domestic service | 297 | 162 | 55% | 1.42 (1.09-1.85) |  | 1.45 (1.10-1.91) |  | 1.44 (1.09-1.90) |  | 1.41 (1.06-1.86) |  |
|  | Prof/tech/man/Cler | 202 | 79 | 39% | 0.76 (0.55-1.04) |  | 0.86 (0.62-1.19) |  | 1.05 (0.73-1.52) |  | 1.07 (0.74-1.54) |  |
|  | Sales & services | 449 | 151 | 34% | 0.60 (0.47-0.76) |  | 0.73 (0.56-0.93) |  | 0.76 (0.58-0.98) |  | 0.76 (0.58-0.98) |  |
|  | Skilled manual | 400 | 138 | 35% | 0.62 (0.49-0.80) |  | 0.78 (0.60-1.01) |  | 0.79 (0.60-1.04) |  | 0.79 (0.60-1.04) |  |
|  | Unskilled manual | 417 | 140 | 34% | 0.60 (0.47-0.76) |  | 0.72 (0.56-0.93) |  | 0.72 (0.55-0.93) |  | 0.72 (0.55-0.94) |  |
|  | Student/pupil | 81 | 22 | 27% | 0.44 (0.27-0.73) |  | 0.86 (0.49-1.51) |  | 1.00 (0.56-1.77) |  | 0.99 (0.55-1.75) |  |
| Religion | Christian | 4129 | 1907 | 46% | Ref | 0.09 | Ref | 0.15 | Ref | 0.24 | Ref | 0.22 |
|  | Islam | 81 | 36 | 44% | 0.93 (0.60-1.45) |  | 0.95 (0.60-1.50) |  | 1.07 (0.67-1.69) |  | 1.06 (0.67-1.69) |  |
|  | Other | 30 | 8 | 27% | 0.42 (0.19-0.95) |  | 0.44 (0.19-1.00) |  | 0.49 (0.21-1.14) |  | 0.48 (0.20-1.11) |  |
|  |  | N | N Attended | % Attended | Unadjusted | p-value | Adjusted for age & sex | p |  |  | Adjusted for everything | p |
| Eye condition | Normal | 209 | 86 | 41% | 0.68 (0.51-0.91) | <0.001 | 0.64 (0.47-0.86) | 0.012 |  |  | 0.66 (0.49-0.89) | 0.008 |
|  | Loss of vision | 1878 | 954 | 51% | Ref |  | Ref |  |  |  | Ref |  |
|  | Chronic | 336 | 158 | 47% | 0.86 (0.68-1.08) |  | 0.85 (0.67-1.08) |  |  |  | 0.84 (0.66-1.07) |  |
|  | Acute | 1368 | 563 | 41% | 0.68 (0.59-0.78) |  | 0.83 (0.71-0.96) |  |  |  | 0.80 (0.68-0.93) |  |
|  | Urgent | 449 | 190 | 42% | 0.71 (0.58-0.87) |  | 0.93 (0.75-1.16) |  |  |  | 0.88 (0.71-1.11) |  |

